# Slider Horizontal Movement and Arm Angle are Associated with Elbow and Forearm Injury Incidence in MLB Pitchers

**DOI:** 10.64898/2026.08.30.26361739

**Authors:** Connor Richards, D. Taylor La Salle, Oscar Vila Dieguez, Samuel R Ward

## Abstract

**Background:** Sweeping sliders with large horizontal break are hypothesized to be associated with arm injury, and prior work suggests a link between slider usage, arm angle, and injury.

**Purpose:** To evaluate whether arm angle and slider horizontal movement are associated with elbow or forearm injury risk in MLB pitchers.

**Study Design:** Retrospective cohort study; Level of evidence, 3

**Methods:** Statcast data from 2020–2025 and injury data were used to study the relationship between sliders and arm injuries. Pitchers with at least 30 innings pitched (IP) were evaluated for same- and next-season injury incidence to the Elbow, Forearm, or Elbow/Forearm. A generalized additive model (GAM) related pitch-level variables to injury incidence, and average marginal effect (AME) odds ratios are reported for main effects.

**Results:** All fits were significant at *p* < .01. Fits for same-season (*N* = 1,957, *p* = .00008, *R*^2^ = .066) and next-season (*N* = 2,511, *p* = .00005, *R*^2^ = .053) Elbow/Forearm injury were significant at *p* < .001. Same-season (*R*^2^ = .061) and next-season (*R*^2^ = .055) Forearm injury fits had *p* = .001. Main effects for slider glove-side movement and fastball usage, their interactions with arm angle, and arm angle main effects were the most predictive features. Across the three fits where the slider glove-side movement main effect was significant, odds ratios ranged from 1.03 to 1.07, meaning that each additional inch of slider glove-side movement was associated with a 3% to 7% increase in the observed injury incidence. Furthermore, this effect was magnified at high arm angle and muted at low arm angle.

**Conclusion:** We observed that slider horizontal movement, arm angle, and their interaction were significantly associated with incidence of Forearm and combined Elbow/Forearm injury. This effect was weak or non-existent at low arm angles and strong at high arm angles, suggesting that arm angle moderates the risk of slider horizontal movement, and providing evidence that sliders with large horizontal break (e.g. sweepers) may pose an injury risk.

## Introduction

The rise of medial elbow injuries among Major League Baseball (MLB) pitchers is one of the most frequently studied and discussed problems in sports medicine today. The literature is replete with analyses of ulnar collateral ligament (UCL) injury risk factors,^2,4,6,9,12^ forearm injury risk factors,^5,14,16,27^ and discussions of the rising rates of injury among MLB pitchers.^7,13,14,22,24^

Puga et al. (2025) found that, notwithstanding a decrease in overall injury rates, MLB has seen a sharp rise in flexor tendon and forearm injuries in the 2023 and 2024 seasons compared to 2021 and 2022.^16^ Moreover, numerous authors have found an empirical link between the incidence of forearm injury and UCL reconstruction (UCLR),^5,27^ and others have argued that more study of the forearm and flexor-pronator mass (FPM) is needed.^14^

Amidst the rise of pitching injuries, MLB pitchers and teams continue to chase “stuff,”^21^ pitching vernacular for pitches with particularly effective movement, which often rely on some combination of velocity, movement, and deception. The sweeper is a new pitch type, a sub-type of slider characterized by extreme horizontal movement towards the pitcher”s glove side and almost no induced vertical movement.^11^ This frisbee-like pitch was first tracked by Statcast in 2023,^11^ with tagging retroactively applied to prior seasons, and its usage more than tripled from 2021 to 2023.^21^ The rise of the sweeper and concomitant rise of arm injuries among MLB pitchers has led to experts to hypothesize that the sweeper drives injury risk, a heretofore not substantiated and altogether contested view.^21^

The incorporation of arm angle data from Statcast may offer an opportunity to evaluate this hypothesis. While a frequent variable in biomechanical analyses, arm slot has been effectively absent from epidemiological studies to date.^15^ One analysis, from Lipa et al. (2025), attempted to measure arm angle for MLB pitchers from game film,^9^ but other approaches have largely ignored arm angle data because it was unavailable from Statcast until the end of the 2024 season.^1^ Recent work has shown that arm angle, slider usage, slider velocity, and their interactions may exhibit a non-linear relationship with injury incidence among MLB pitchers.^19^

We propose to evaluate the relationship between slider horizontal movement and arm angle with elbow and forearm injury among MLB pitchers by modeling the relationship between slider usage, horizontal movement, velocity, and other pitch-level variables, including interaction effects, and incidence of elbow or forearm injury from MLB injured list (IL) data.

## Methods

Pitch-level data from Statcast was accessed for MLB games spanning the 2020 to 2025 seasons using pybaseball,^8^ and injured list data was extracted from transactions obtained via the MLB Stats API.^10^ Statistical analysis and modeling were performed using R (Version 4.6.0)^18^ and the mgcv package^26^ using α = .05. Data was retrieved and visualized using Python (Version 3.11.14).^17^

### Injury categorization and inclusion criteria

Placements on the MLB injured list were used to identify elbow and forearm injuries to pitchers. Transactions obtained via the MLB Stats API^10^ were categorized based on whether they referenced the elbow or forearm using a list of keywords.^19^ Confirmed UCL reconstruction cases were identified using a public database of UCLR and used to tag elbow injuries to supplement IL placement data for cases where UCLR immediately followed forearm injury.^19,20^

For a season to be included, a pitcher was required to throw at least one fastball (FF, SI, or FC) and face at least 45 hitters, allowing pitch tracking metrics to stabilize. Pitchers were also required to throw at least 30 innings the season preceding injury.

When identifying next-season injury, pitchers were not required to have pitched that season. To verify that IL placements without any IP requirement to identify next-season injury did not introduce selection bias, results were compared to pitchers who faced at least 45 hitters.

### Statcast variables

Statcast reports pitcher arm angle as the angle between the shoulder and hand at ball release, normalized such that 0° is a sidearm release with the arm parallel to the horizontal. This definition, unlike some definitions of arm slot, is irrespective of the angle of the humerus relative to the trunk. This means that a pitcher can achieve higher or lower Statcast arm angle through various means which may be unrelated to glenohumeral abduction. For example, a pitcher may achieve higher arm angle through contralateral trunk tilt without any change in glenohumeral abduction. In addition to arm angle, each pitch has a recorded velocity, spin rate, horizontal movement, and induced vertical movement, among other features.

Data was aggregated across five pitch types representing either fastballs or sliders. Four-seam fastballs (FF), sinkers (SI), and cutters (FC) were categorized as fastballs. Sliders (SL) and sweepers (ST) were categorized as sliders. All non-breaking ball and non-fastball pitches were categorized as off-speed and grouped together, with the goal of capturing pitchers” usage of changeups (CH), splitters (FS), and other offspeed offerings. Velocity, spin rate, and usage were computed for each of fastballs, sliders, and offspeed pitches. Glove-side movement, the horizontal movement towards the pitcher”s glove side, was also computed for sliders.

The Statcast designation of “sweeper” identifies a slider with increased horizontal break.^11^ Notwithstanding hypotheses about the sweeper as a potential driver of injury, the pitch itself is still relatively infrequently used.^21^ As a result, requiring pitchers to throw a sweeper would needlessly cut down the available sample size. Instead, we used the average horizontal break of all the sliders a pitcher threw, measured as the glove-side movement, as a proxy for sweeper usage and sweeper movement to evaluate the relationship to injury. As a check on the hypothesis that slider horizontal movement would capture the rise in sweeper usage, the change in slider horizontal break from 2020 to 2025 was also evaluated using a linear mixed model.

### Generalized additive model (GAM) fit

Features were evaluated for inclusion using a bivariate screen for injury type (Elbow, Forearm, Elbow/Forearm) and timeline (same season, next season). Features with *p* < .20 were selected for inclusion in the final model. Given that a pitcher may appear in multiple seasons from 2020 to 2025, we evaluated mixed effects in addition to season-level variables.

The permissive filter (*p* < .20) was used to select variables for inclusion in conjunction with a reduced maximum likelihood (REML) optimization method as another automated feature selection step at the time of model fitting. This choice allowed many pitch variables and tensor product over those variables to be screened for their potential contribution to the model while controlling against overfitting. Variables were modeled as cubic splines with basis dimension 4 and included as spline smooths over a single variable or tensor product over two variables including interactions. For interaction terms, the full tensor product, inclusive of variable main effects plus interactions (*te* in mgcv), was screened for inclusion. Interactions selected for inclusion were decomposed as the sum of main effects (*s* in mgcv) plus marginal interaction (*ti* in mgcv) in the final fit. For example, *te*(*arm angle, fastball usage*) was decomposed to *s*(*arm angle*) + *s*(*fastball usage*) + *ti*(*arm angle, fastball usage*).

Nominal degrees of freedom, effective degrees of freedom, (*n*_*dof*_), fit- and feature-level p-values, and Nagelkerke pseudo-*R*^2^ values are presented for each fit.

## Results

N = 1,957 pitcher-seasons met inclusion criteria for same-season injury risk, and N = 2,511 pitcher-seasons met inclusion criteria for next-season injury risk. We found slider horizontal movement increased year-over-year from 2020 through 2025, in line with the rise in sweeper usage during that period,^21^ which is illustrated in *Figure 1b*. The average pitcher in our dataset gained 0.26 inches of glove-side movement slider movement per year from 2020 to 2025 (*p* < 10^−5^), while no significant trend was observed for the change in slider velocity at the pitcher-level (*p* = .61).

**Figure 1:**
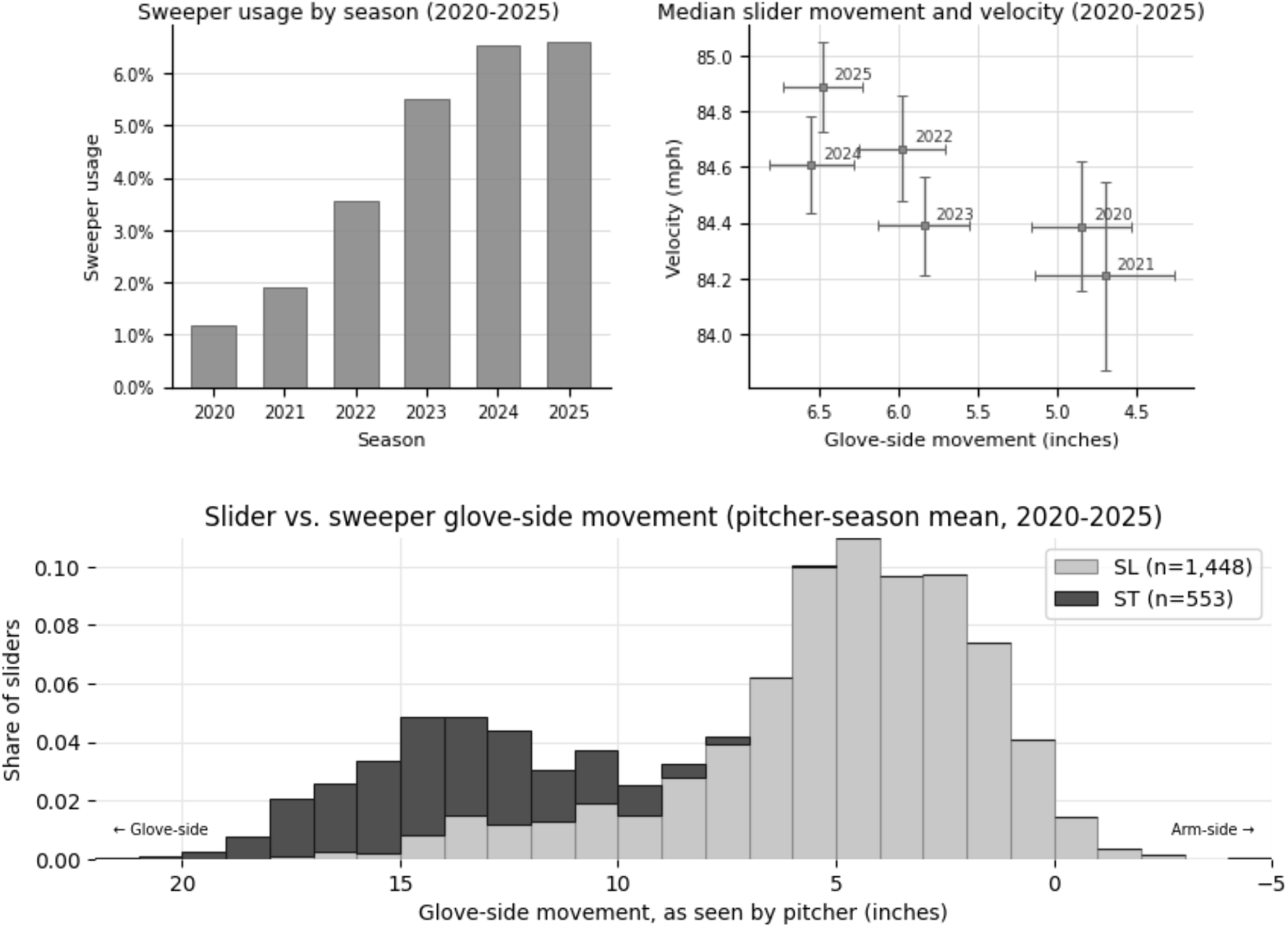
Sweeper usage and pitch shapes among included pitchers, 2020 to 2025. (a, left) Cohort-wide sweeper usage by season as a share of total pitches thrown. (b, right) Median slider (SL + ST) velocity (mph) and glove-side break (inches). Increase in slider glove-side movement was statistically significant (p < 10^−5^) using a linear mixed model. (c, bottom) Mean glove-side break by pitcher season for sliders (SL) and sweepers (ST). Note: axes oriented to reflect glove-side movement increasing to the left.

For injury fits, mixed effects were not significant in any models in which they were selected. On average, mixed effect fits achieved higher *R*^2^ values at the cost of higher effective degrees of freedom, and the features identified in those fits were consistent with those using season-level pitch data alone. In what follows, we omit mixed effects and report season-level fits in the interest of simplicity and limiting effective degrees of freedom.

### Slider and sweeper shapes

*Figure 1c* shows the distribution of glove-side movement for pitches tagged as sliders (SL) and sweepers (ST). Taken together, the distribution of slider horizontal break, with mass near 5 inches of glove-side movement corresponding to Statcast-tagged “sliders” and near 15 inches of corresponding to Statcast-tagged “sweepers.” The substantial low tail of the tagged “slider” distribution meant there was non-trivial overlap between “sliders” and “sweepers” across pitchers. Combining any sliders or sweepers thrown by a given pitcher allowed us to use glove-side movement as a proxy for sweeper usage and evaluate the relationship between slider horizontal movement, injury, and other pitch variables.

### Fit results

The choice of *p* < .20 as a threshold for inclusion was a very permissive initial variable screen. Of the 21 variables screened per model, between 13 and 17 were chosen across the different fits. The reduced maximum likelihood fitting method was instead a much more stringent feature selection method, taking the large volume of variables provided and eliminating between 91% and 96% of the nominal degrees of freedom available to each model. A total of 15 features had *n*_*dof*_ > 0.01 in at least one fit, and 11 features were significant in at least one fit. All six fits were statistically significant, and *R*^2^ values were largely consistent across same-season 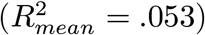 and next-season injury prediction 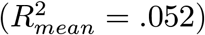.

Arm angle (*n*_*dof*_ = 6.1, *n*_*fits*_ = 5), fastball usage (*n*_*dof*_ = 5.2, *n*_*fits*_ = 4), slider glove-side break (*n*_*dof*_ = 3.0, *n*_*fits*_ = 3), and slider velocity (*n*_*dof*_ = 3.0, *n*_*fits*_ = 2) were the only main effect terms significant in more than one fit. The marginal interaction effect of arm angle with fastball usage (*n*_*dof*_ = 6.4, *n*_*fits*_ = 4) and with slider glove-side break (*n*_*dof*_ = 3.6, *n*_*fits*_ = 3) were the only interactions significant in more than one fit.

### Slider horizontal break and velocity

Consistently across fits for Forearm injury and Elbow/Forearm injury, increased slider movement at high arm angles was associated with increased incidence of injury, and one or both of slider glove-side movement and its interaction with arm angle were significant in all four fits. However, in both same-season and next-season Elbow injury fits, no relationship was observed between slider horizontal movement and injury incidence, as feature selection penalized the contribution of both main effect and interaction terms to 0. For Forearm and Elbow/Forearm fits, injury incidence increased as a function of both arm angle and horizontal break while holding the other constant, but the strength of this relationship varied as a function of arm angle and slider movement.

**Table 1:** Feature-level p-values for bivariate screens. Bold denotes features selected for multivariate fits; asterisks denote significance, * < .05, ** < .01, *** < .001.

| Feature | Elbow Same Year | Forearm Same Year | Elbow or Forearm Same Year | Elbow Next Year | Forearm Next Year | Elbow or Forearm Next Year |
| --- | --- | --- | --- | --- | --- | --- |
| Arm Angle | **0.071** | **0.004\*\*** | **0.001\*\*** | **0.026\*** | **0.034\*** | **0.002\*\*** |
| Fastball Velocity | 0.826 | 0.217 | 0.298 | **0.119** | 0.288 | **0.055** |
| Fastball Usage | **0.192** | **0.150** | 0.209 | **0.184** | **0.085** | **0.040\*** |
| Fastball Spin Rate | 0.594 | 0.204 | 0.275 | **0.089** | 0.571 | **0.124** |
| Slider Velocity | 0.206 | **0.043\*** | **0.024\*** | **0.034\*** | **0.059** | **0.008\*\*** |
| Slider Usage | **0.042\*** | 0.902 | **0.114** | **0.014\*** | 0.480 | **0.017\*** |
| Slider Spin Rate | 0.870 | **0.021\*** | **0.150** | 0.618 | **0.166** | 0.243 |
| Fastball Velocity × Arm Angle | 0.202 | **0.021\*** | **0.009\*\*** | **0.050** | **0.008\*\*** | **0.003\*\*** |
| Fastball Usage × Arm Angle | **0.017\*** | **0.004\*\*** | **0.0003\*\*\*** | **0.034\*** | **0.013\*** | **0.003\*\*** |
| Fastball Spin Rate × Arm Angle | **0.038\*** | **0.010\*\*** | **0.001\*\*** | **0.022\*** | **0.026\*** | **0.004\*\*** |
| Slider Velocity × Arm Angle | **0.033\*** | **0.019\*** | **0.004\*\*** | **0.023\*** | **0.042\*** | **0.002\*\*** |
| Slider Usage × Arm Angle | **0.023\*** | **0.013\*** | **0.002\*\*** | **0.003\*\*** | 0.055 | **0.0003\*\*\*** |
| Slider Spin Rate × Arm Angle | **0.039\*** | **0.004\*\*** | **0.003\*\*** | **0.021\*** | **0.069** | **0.002\*\*** |
| Offspeed Spin Rate × Arm Angle | **0.027\*** | **0.047\*** | **0.005\*\*** | **0.010\*** | **0.017\*** | **0.003\*\*** |
| Offspeed Usage × Arm Angle | **0.185** | **0.028\*** | **0.008\*\*** | **0.059** | **0.024\*** | **0.001\*\*** |
| Offspeed Velocity × Arm Angle | **0.078** | **0.032\*** | **0.007\*\*** | **0.079** | **0.061** | **0.007\*\*** |
| Slider Glove-Side Break × Arm Angle | **0.055** | **0.014\*** | **0.007\*\*** | **0.029\*** | **0.149** | **0.007\*\*** |
| Offspeed Spin Rate | **0.074** | 0.990 | **0.096** | **0.013\*** | **0.108** | 0.263 |
| Offspeed Usage | 0.201 | 0.848 | **0.106** | 0.290 | 0.367 | 0.707 |
| Offspeed Velocity | 0.749 | 0.942 | 0.634 | 0.287 | 0.313 | **0.170** |
| Slider Glove-Side Break | 0.772 | **0.146** | 0.394 | 0.765 | 0.370 | 0.731 |

**Table 2:**
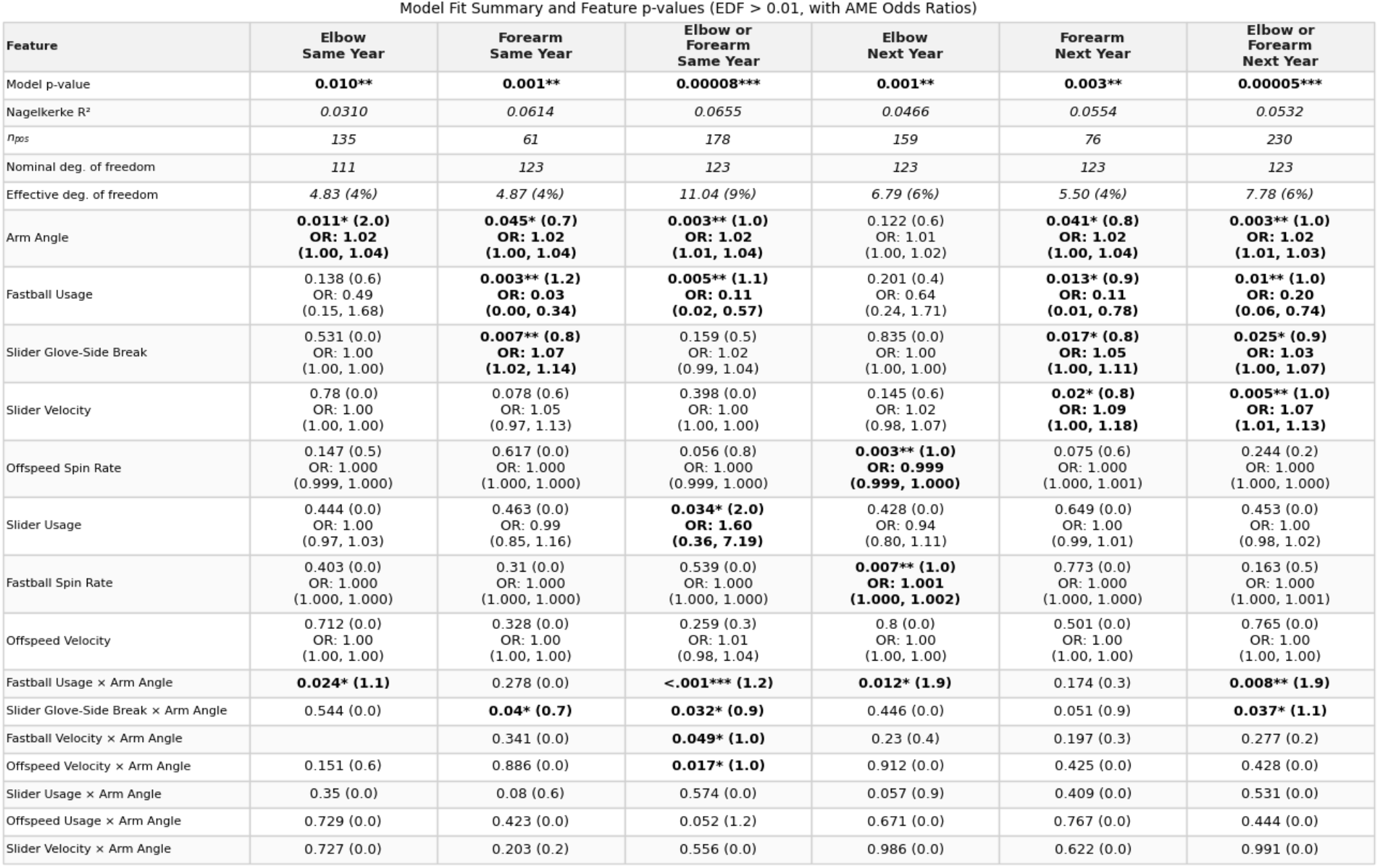
Fit summaries and term-level p-values for GAM fits. Model-level effective degrees of freedom displays n_dof_ and the share of nominal degrees of freedom used by the model. Interaction effect rows show p-value and n_dof_ and are ordered by total n_dof_ across all models. Main effect rows also include AME odds ratios. Filtered to features with n_dof_ > .01 in at least one model. Asterisks denote significance, * < .05, ** < .01, *** < .001.

For next-season Elbow/Forearm injury, average marginal effect (AME) odds ratios for slider horizontal movement varied across arm angle quartiles. For the first quartile 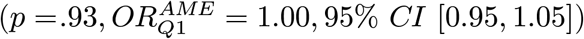 and second quartile 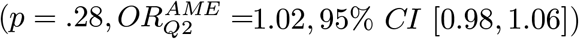 of arm angle, odds ratios for slider horizontal break were consistent with unity. Among pitchers in the third quartile by arm angle, the average marginal effect of slider glove-side movement showed a statistically significant association with higher incidence of injury 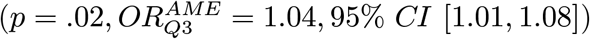. The effect was larger and statistically significant for the fourth quartile 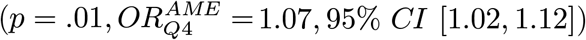, where a 1 inch increase in slider horizontal movement increased odds of injury incidence by 7%. *Figure 2a* shows the dependence of Elbow/Forearm injury incidence on arm angle and slider glove-side movement.

**Figure 2:**
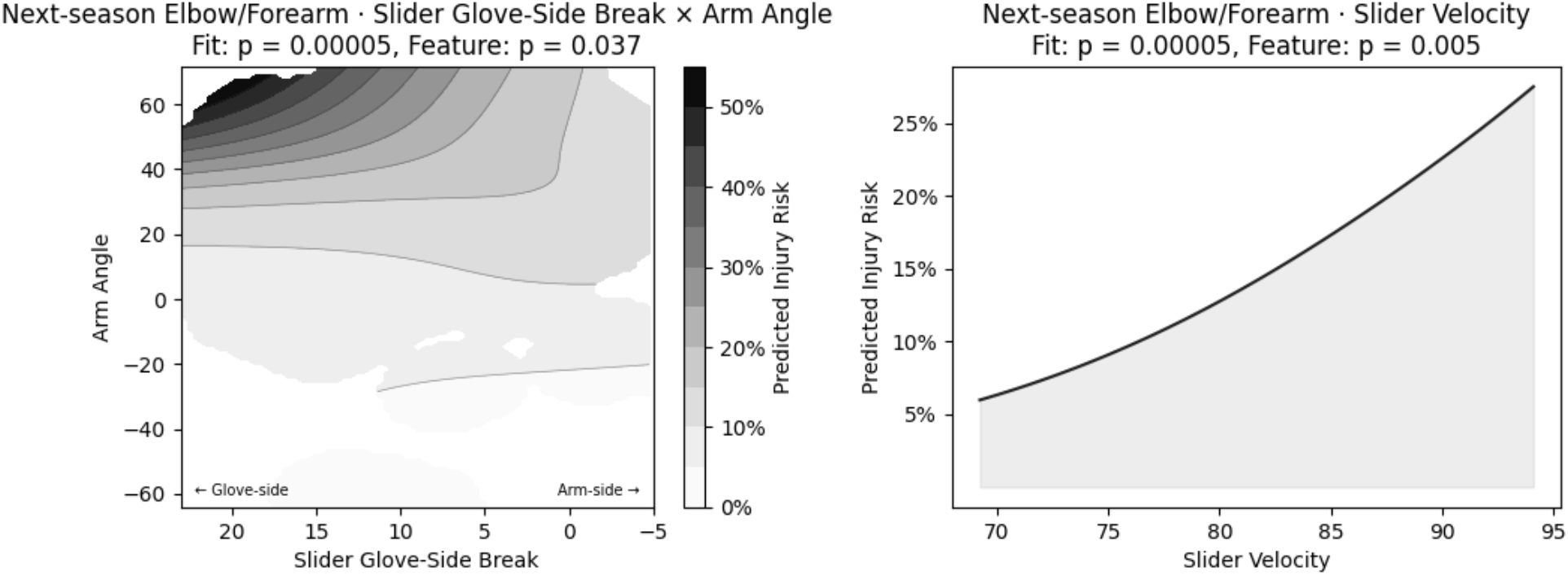
Non-plotting variables held constant at cohort medians. (a) Modeled next-season Elbow/Forearm injury probability as a function of arm angle and slider horizontal break; note that x-axis sign is flipped so that left is positive. (b) Modeled next-season Elbow/Forearm injury probability as a function of slider velocity.

Similarly, at low values of slider glove-side movement, the relationship between arm angle and injury incidence was weaker, increasing markedly at high values of slider movement. The average marginal effect of arm angle on next-season Elbow/Forearm injury among pitchers in the first 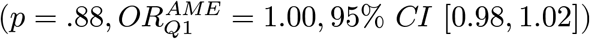 and second 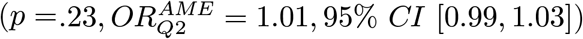 quartiles of slider glove-side movement were not significant. The effect was significant for the third 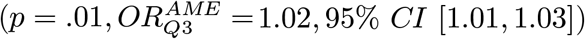 and fourth quartile 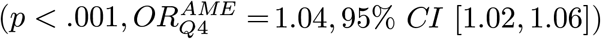 by slider horizontal movement, where a 1° increase in arm angle was associated with a 2% and 4% increase in next-season Elbow/Forearm injury incidence, respectively.

Slider velocity and slider glove-side movement main effects were also associated with increased incidence of next-season Forearm and Elbow/Forearm injury. Slider glove-side movement was also associated with increased incidence of same-season Forearm injury 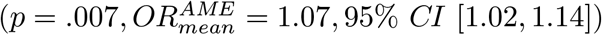, where a 1 inch increase in slider horizontal movement was associated with a 7% increase in Forearm injury incidence.

### Fastball usage

Fastball usage also significantly predicted injury incidence. It was significantly, inversely related to injury incidence for same- and next-season fits of Forearm and Elbow/Forearm injury. Reducing fastball usage here is equivalent to increasing usage of breaking balls and offspeed pitches, meaning that this result could be interpreted to say that increasing non-fastball usage is associated with increased injury incidence. Average marginal effect odds ratios for these four fits ranged from . 03 to . 20. Equivalently stated per 10% change in fastball usage, this range is. 70 to . 85, meaning that a 10% increase in fastball usage was associated with a reduction in injury incidence ranging from 30% to 15%, respectively. Main effects were not significant for same-season or next-season Elbow injury.

The marginal interaction between fastball usage and arm angle significantly predicted same-season (*p* = .02) and next-season Elbow (*p* = .01) injury and same-season (*p* < .001) and next-season Elbow/Forearm injury (*p* = .01). At low arm angles, increased fastball usage is generally associated with reduced injury incidence. However, at higher arm angles, that effect was often muted.

### Fastball and offspeed velocity

The only fit in which a term containing fastball or offspeed velocity was significant was same-season Elbow/Forearm injury, where the marginal interaction of both variables with arm angle was significant. The main regions of elevated injury incidence were in pitchers who threw at high arm angle and low fastball velocity, as well as high arm angle and high offspeed velocity, as shown in *Figure 3*.

**Figure 3:**
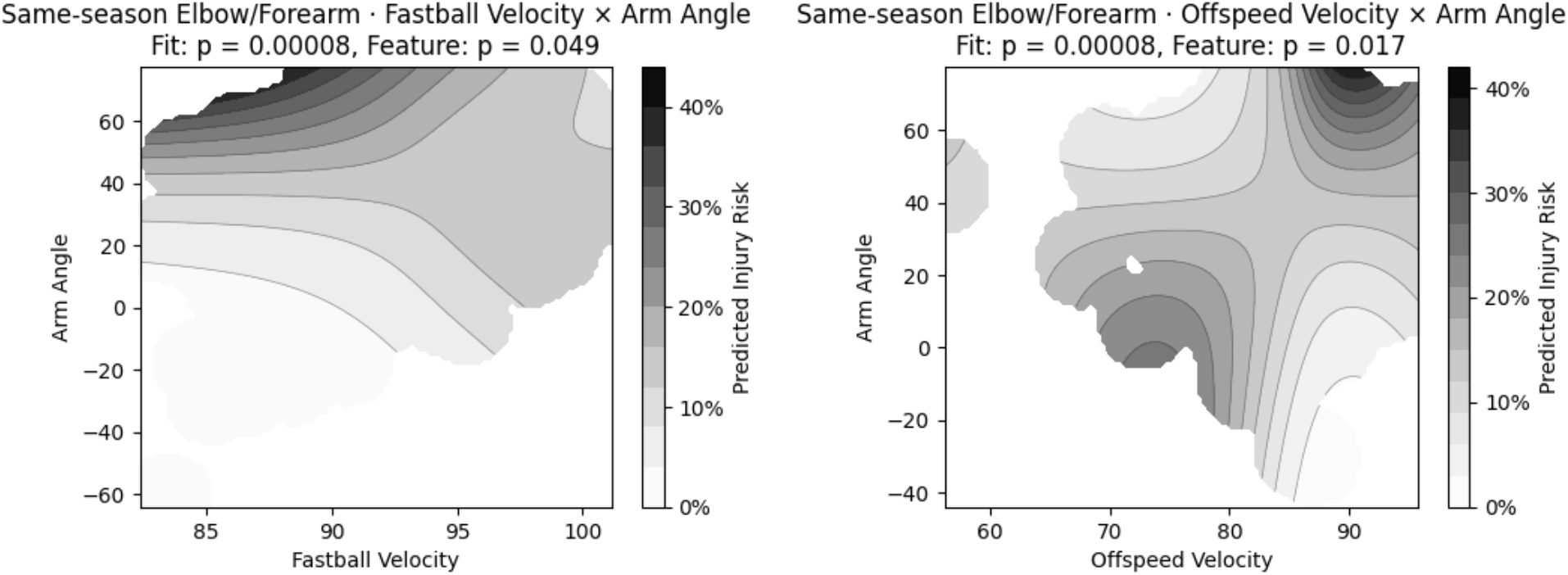
Non-plotting variables held constant at cohort medians. Modeled same-season Elbow/Forearm injury probability as a function of (a) arm angle and fastball velocity and (b) arm angle and offspeed velocity.

### Spin rate

Two countervailing effects were observed for spin rate and next-season Elbow injury. Fastball spin rate was positively related to injury incidence 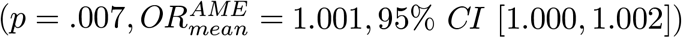, while offspeed spin rate was negatively associated with injury 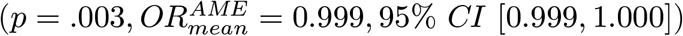. These were the only two terms, main effect or interaction, containing spin rate which were significant in any fit.

### Next-season injury fit

When fitting and reporting next-season injury, we did not require any minimum number of innings pitched or batters faced in the “next season” during which injury was observed from injured list placements. To verify the validity of this approach, we performed fits for pitchers who faced at least 45 hitters during the season in which they were injured.

Elbow (*p* = .001, *n*_*pos*_ = 136, *R*^2^ = .054), Forearm (*p* = .003, *n*_*pos*_ = 62, *R*^2^ = .069), and Elbow/Forearm (*p* < .001, *n*_*pos*_ = 180, *R*^2^ = .067) fits were all significant, and similar variables were identified as being associated with incidence of injury.

## Discussion

We found that the relationship between slider horizontal movement and injury incidence is moderated by arm angle among MLB pitchers. Slider horizontal movement, arm angle, and their marginal interaction were significant predictors in both same-season and next-season fits for Elbow/Forearm and Forearm injury. Fastball usage, whose interaction with arm angle has been previously identified as a risk factor,^19^ remained an important predictor of injury, as did slider velocity.

In addition to the relationship observed between injury incidence, we also found a statistically significant increase in slider horizontal movement year-over-year from 2020 through 2025. These two findings may offer insight into one factor affecting injury risk among MLB pitchers when taken together with the recent rise in forearm flexor injuries.^16^ As hypothesized previously, there does appear to be a link between sliders with pronounced glove-side movement and arm injury incidence.^21^ Furthermore, there is evidence to suggest this association may be even stronger when those sliders are thrown at high arm angle. The extent to which this effect is moderated by arm angle may explain why observational evidence of this relationship had previously remained elusive.

Pseudo-*R*^2^ values for the fits presented ranged from . 031 for next-season Elbow injury to . 066 for same-season Elbow/Forearm injury. These values are in line with performance reported by Chalmers et al. (2016) using pitch tracking variables, who achieved Nagelkerke pseudo-*R*^2^ of . 049 when predicting UCLR using pitch tracking variables.^2^ The inclusion of slider horizontal movement and offspeed variables also improved on prior work which used arm angle and other pitch tracking variables to predict same-season Elbow injury (*R*^2^ = .019), Forearm injury (*R*^2^ = .052), and Elbow/Forearm injury (*R*^2^ = .043) by roughly . 01 per fit on average.^19^

Furthermore, while our analysis does not directly evaluate a mechanism by which arm angle could moderate the relationship between slider horizontal movement and arm injury, one of the results presented here may suggest a candidate. Notably, we found that throwing sliders with extreme glove-side movement was associated with forearm-only and elbow or forearm injury incidence, but we did not observe a relationship with elbow-only injury incidence. The association with forearm injury aligns with the hypothesis that sweepers may place additional load on the flexor-pronator mass (FPM) rather than the UCL, which would explain the observed rise in forearm injuries amidst the increases in sweepers and slider glove-side movement more generally.^5,16,21^ Additionally, the nature of the mechanics required to achieve a sweeper, a supinated release that generates a vertical spin axis to produce frisbee-like movement, requires recruitment of the FPM to move the forearm at a time when muscles like the flexor carpi ulnaris and others are performing their role of stabilizing the medial elbow against valgus load.^3,23,25^ If the brunt of the load placed on the arm by sweepers is borne by the FPM, this would offer an explanation for the rise in forearm injuries, the observational link between slider glove-side movement and injury observed here, and an explanation of the role that slider and offspeed usage plays in understanding injury risk: the FPM, when hampered due to injury or the demands of throwing a sweeper, may struggle to adequately stabilize the UCL.

Some of the tension in the literature related to sweeper usage as a risk factor for injury arose because anecdotal evidence about the risk posed by sweepers appeared to clash findings indicating that sliders and breaking balls were not any more stressful than fastballs.^21^ This finding suggests biomechanical analyses of the stress placed on the arm by fastballs and sliders may benefit from studying the potentially moderating effect of arm angle. This is especially true as it relates to the ability of the FPM to effectively stabilize the medial elbow under the increased load of greater abduction and external rotation at higher arm angles, and future work should explore this.

It should be noted that our analysis here is not without limitations. We evaluated the relationship between pitch-tracking variables and placement on the injured list, which is a useful and publicly available proxy for injury but is susceptible to numerous potential shortcomings.^19^ Furthermore, in an effort to study the impact of pitch shapes, we did not include powerful predictors like history of UCL reconstruction or workload variables, nor do we evaluate here the temporal interrelationship of elbow or forearm injury, changing arm angle, and their effect on pitch shapes. Instead, we attempted to simply evaluate pitch shape-based associations with injury incidence, and future work is needed to understand the extent to which these provide meaningful signal for predictive models of future injury over and above injury history, workload, age, and other features.

Additionally, while we found empirical evidence linking arm angle, pitcher arm injury, and horizontal movement on sliders, we do not directly model sweeper usage as a risk factor. Horizontal slider movement was instead used as an accessible proxy for sweeper usage. Given the limited, albeit growing, usage of sweepers among MLB pitchers and the higher usage of sliders, together with the shape variability within pitch types, the choice to use glove-side movement on the slider is both analytically convenient and potentially encodes additional information that sweeper usage or slider usage alone may not. As *Figure 1c* shows, some pitchers throw “sliders” with more horizontal movement than others “sweepers”, and each pitch subtype has substantial variability in observed horizontal movement, meaning that the average horizontal movement averaged across sliders and sweepers captures additional information about the specifics of each pitcher”s respective shapes. Future work should look to evaluate the extent to which individual pitch shapes on the slider and sweeper, their respective usage, and arm angle are associated with arm injury, especially as more data becomes available.

## Conclusion

Slider horizontal movement and its interaction with arm angle were significantly associated with incidence of elbow and forearm injury among MLB pitchers from 2020 to 2025. These effects were observed in both the Forearm and combined Elbow/Forearm fits, but they were not significant for predicting Elbow injury alone. Arm angle, fastball usage, and their interaction were also found to be significantly associated with injury incidence.

While others have hypothesized the link between sweeping sliders and injury,^21^ to the best of the authors” knowledge, this is one of the first analyses linking sliders with large horizontal movement, like the sweeper, to injury incidence and identifying arm angle as a variable which moderates this relationship. Pseudo-*R*^2^ values of . 031 to . 066 meet or exceed the performance of existing estimates of elbow injury incidence using pitch tracking variables, indicating that the inclusion of slider horizontal movement and its interaction with arm angle provide meaningful signal related to injury incidence.

Furthermore, the fact that this remains a strong predictor for next-season Forearm and Elbow/Forearm injury incidence suggests that the effect observed is not simply explained by concomitant changes to a pitcher”s slider or arm angle resulting from injury. Instead, arm angle and slider horizontal movement, along with fastball usage and slider velocity, are all significantly associated with injuries to the elbow or forearm in the following season.

## Data Availability

All input data for the present study are publicly available online at their respective sources. Engineered data from the analysis performed may be made available upon reasonable request to the authors.

https://baseballsavant.mlb.com

https://statsapi.mlb.com

https://docs.google.com/spreadsheets/d/1gQujXQQGOVNaiuwSN680Hq-FDVsCwvN-3AazykOBON0

